# Participant Experience with the SpaceLabs 90227 ABPM and SOMNOmedics ABPM Pro Devices

**DOI:** 10.64898/2026.07.27.26359028

**Authors:** Josephine Soddano, Brandon Fernandez-Sedano, Sumayya Shurovi, Michelle L. David, Guixiao Ding, Fatma Dansoko, Joseph E. Schwartz, Marwah Abdalla

**Author notes:** **Corresponding Author:** Marwah Abdalla, MD, MPH, 622 West 168^th^ Street, Columbia University Medical Center, New York, NY 10032. **Author Disclosures:** No relevant financial disclosures.

## Abstract

Ambulatory blood pressure monitoring (ABPM) is recommended for confirming hypertension and assessing out-of-office blood pressure (BP). However, patient burden and device tolerability may limit broader implementation. We compared participant experience with a traditional oscillometric ABPM device and a compact cuff-integrated ABPM.

The PRO-BP Study was a pilot randomized crossover study of 20 adults in New York City. Participants completed two 24-hour ABPM periods over 7 days using the SpaceLabs 90227 and SOMNOmedics ABPM Pro devices. After each period, participants rated comfort, pain, sleep interference, embarrassment, noise, skin irritation, and interference with daytime activities.

Both devices achieved guideline-based recording-quality thresholds. Compared with SpaceLabs, ABPM Pro was associated with greater comfort (median 7.0 [IQR, 5.0-8.5] vs 3.5 [IQR, 2.0-6.5]; P=0.004), less pain (1.5 [0-3.5] vs 5.0 [0.5-7.0]; P=0.003), and less embarrassment (0.5 [0-3.5] vs 3.0 [0-6.0]; P=0.01). Other experience ratings did not differ significantly.

Participant experience should be considered alongside recording quality when evaluating validated ambulatory BP monitoring technologies.

## Introduction

Although ambulatory blood pressure monitoring (ABPM) is guideline recommended for confirming hypertension and characterizing out-of-office BP phenotypes,^1^ patient burden remains an important barrier to broader adoption.^2-4^ Participants report sleep disturbance during ABPM, with additional concerns related to discomfort, device bulkiness, and embarrassment while wearing the monitor in public.^4-7^

Recent advances in BP monitoring technology have introduced novel devices designed to improve patient experience. The SOMNOmedics ABPM Pro is a validated, FDA-cleared, upper-arm oscillometric ABPM device with a compact, tubeless, cuff-integrated design.^8,9^ Evaluating the acceptability of emerging validated BP technologies has been identified as a research priority in a recent National Heart, Lung, and Blood Institute (NHLBI) report.^2^ We conducted a pilot within-person randomized crossover study comparing the SpaceLabs ABPM device with the SOMNOmedics ABPM Pro device to evaluate participant tolerability and experience.

## Methods

### Study Design

The PRO-BP Study was a pilot randomized crossover study comparing participant experience between the SpaceLabs 90227 ABPM (SpaceLabs Healthcare, Snoqualmie, WA, USA) and the SOMNOmedics ABPM Pro device (SOMNOmedics GmbH, Randersacker, Germany). Participants completed three study visits and two separate 24-hour ambulatory BP monitoring periods during a 7-day monitoring period (**Supplemental Figure 1**). Detailed study procedures are in the **Supplemental Methods**. The study was approved by the Columbia University Irving Medical Center Institutional Review Board and all participants provided written informed consent.

Participants were randomized (1:1) to one of two monitoring sequences: SpaceLabs followed by ABPM Pro or ABPM Pro followed by SpaceLabs. The SpaceLabs 90227 is a validated oscillometric ABPM device widely used in clinical practice and research (Figure 1a).^10^ The SOMNOmedics ABPM Pro is a compact tubeless oscillometric ABPM device (Figure 1b).^8,9^ Both devices were fitted on the non-dominant arm using an appropriately sized cuff and programmed to measure BP every 30 minutes. Sleep and wake periods were defined using wrist actigraphy supplemented by sleep diaries.

**Figure 1.**
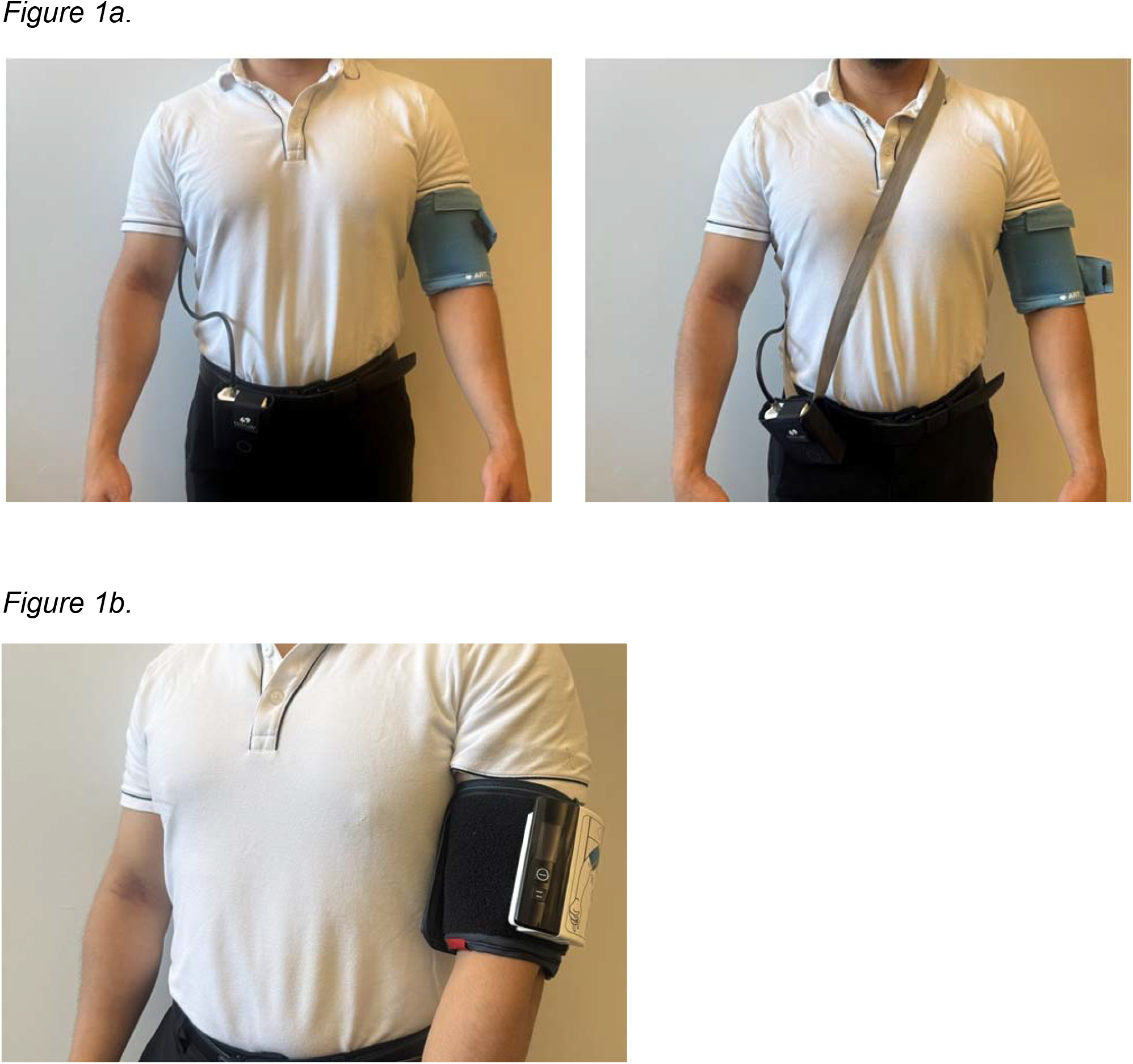
Ambulatory blood pressure monitoring devices used in the PRO-BP Study. **(A)** SpaceLabs 90227 device consisting of an upper-arm cuff connected by tubing to a monitor worn on the waist or shoulder strap. **(B)** SOMNOmedics ABPM Pro device, a compact tubeless ambulatory blood pressure monitor attached directly to the upper-arm cuff.

### Study Outcomes

Recording quality was evaluated using the 2021 European Society of Hypertension (ESH) and 2024 European Society of Cardiology (ESC) hypertension guideline recommendations.^11,12^ We assessed the proportion of participants meeting ESH (≥20 valid awake and ≥7 valid sleep readings)^11^ and ESC (≥27 valid 24-hour readings) recording quality criteria.^12^ Mean awake and sleep systolic and diastolic BP were calculated using all available readings obtained during the respective actigraphy-defined time periods. Mean 24-hour BP was calculated as the weighted mean of awake and sleep BP values, with weights equal to the proportion of the monitoring period spent awake and asleep.^13^ All participants completed both ABPM monitoring periods and actigraphy and were included in the analysis.

After each monitoring period, participants completed a device-experience questionnaire rating comfort, pain, sleep interference, embarrassment, noise, skin irritation, and interference with daytime activities using a 10-point Likert scale.^14^

### Statistical Analysis

Participant characteristics are presented as mean ± standard deviation (SD) or median (interquartile range [IQR]), as appropriate. Questionnaire responses were compared using Wilcoxon signed-rank tests. Paired comparisons of ambulatory BP measurements between devices were performed using paired t-tests in exploratory analyses. Recording quality metrics, including the proportions of participants meeting ESH and ESC criteria were compared according to monitoring period (Visit 1 vs. Visit 2) using exact McNemar tests. Analyses were performed using R version 4.5.2. A two-sided P value <0.05 was considered statistically significant.

## Results

### Participant Characteristics

Of 26 screened individuals, 23 were eligible, and 20 completed the consent process and both ABPM periods. Median age was 46 years (IQR, 29-51), 45% were male, 45% had a prior clinician diagnosis of hypertension, and 15% used antihypertensive medication. Additional participant characteristics are shown in **Table 1**. Office BP did not differ significantly between Visit 1 and Visit 2 (**Supplemental Results**).

**Table 1.** Participant Characteristics.

| **Characteristic** | **N = 20** |
| --- | --- |
| Age, years | 46 (29–51) |
| Male sex, % | 45 |
| Race/Ethnicity |  |
| White, % | 35 |
| Black, % | 25 |
| Asian, % | 25 |
| Other/Unknown, % | 15 |
| Hispanic ethnicity, % | 15 |
| Height, cm | 171 (166–176) |
| Weight, kg | 75.5 (65.0–88.9) |
| BMI, kg/m<sup>2</sup> | 25.7 (23.8–29.3) |
| College graduate or higher, % | 65 |
| Current smoking, % | 10 |
| Prior history of diabetes, % | 5 |
| Prior history of hypertension, % | 45 |
| Antihypertensive medication use, % | 15 |
| Office hypertension, % | 55 |
| History of obstructive sleep apnea, % | 25 |
| Sleep medication use, % | 10 |
| Insomnia symptoms (ISI ≥15), % | 5 |
| PSQI score | 4.3 ± (2.8) |
| Poor sleep quality (PSQI >5), % | 35 |

Data are presented as percentage, median (interquartile range), or mean ± standard deviation. Abbreviations: BMI, body mass index; ISI, Insomnia Severity Index; PSQI, Pittsburgh Sleep Quality Index.

### Recording Quality and Comparison of BP Measurements

Recording quality was high with both devices (**Table 2**). Median numbers of valid awake readings were 30 (IQR, 27–33) for the SpaceLabs device and 26 (IQR, 25–27) for the ABPM Pro device. Median numbers of valid sleep readings were 13 (IQR, 12– 15) and 14 (IQR, 12–16), respectively Median numbers of valid 24-hour readings were 44 (IQR, 40–46) and 40 (IQR, 37–44), respectively. Nineteen of 20 participants (95%) met the predefined criteria of ≥20 valid awake readings with both devices.

**Table 2.** Ambulatory Blood Pressure Monitoring Recording Quality.

| **Metric** | **SpaceLabs ABPM**<br>**(N = 20)** | **SOMNOmedics ABPM Pro**<br>**(N = 20)** |
| --- | --- | --- |
| Attempted awake readings, median (IQR) | 45 (42–55) | 41 (34–46) |
| Valid awake readings, median (IQR) | 30 (27–33) | 26 (25–27) |
| Participants with $\geq 20$ valid awake BP readings, n (%)<sup>□</sup> | 19 (95) | 19 (95) |
| Attempted sleep readings, median (IQR) | 15 (12–18) | 15 (14–17) |
| Valid sleep readings, median (IQR) | 13 (12–15) | 14 (12–16) |
| Participants with $\geq 7$ valid sleep BP readings, n (%)<sup>□</sup> | 19 (95) | 20 (100) |
| Attempted 24-hour readings, median (IQR) | 61 (58–70) | 58 (49–60) |
| Valid 24-hour readings, median (IQR) | 44 (40–46) | 40 (37–44) |
| Participants with $\geq 27$ valid 24-hour BP readings, n (%)<sup>□</sup> | 19 (95) | 20 (100) |

Abbreviations: ABPM, ambulatory blood pressure monitoring; IQR, interquartile range.
□ Defined as $\geq 20$ valid awake BP readings, consistent with the 2021 European Society of Hypertension guideline recommendations.
□ Defined as $\geq 7$ valid sleep BP readings, consistent with the 2021 European Society of Hypertension guideline recommendations.
□ Defined as $\geq 27$ valid 24-hour BP readings, consistent with the 2024 European Society of Cardiology guideline recommendations.

Corresponding proportions achieving ≥7 valid sleep BP readings were 19 (95%) and 20 (100%), respectively. Nineteen participants (95%) wearing the SpaceLabs device and all 20 participants (100%) wearing the ABPM Pro device met the 2024 ESC criteria of ≥27 valid 24-hour readings. Recording quality did not differ between the first and second ABPM recording periods for the awake, sleep, or 24-hour recording quality metrics (all P values > 0.05). Exploratory comparisons of ambulatory BP measurements demonstrated higher awake and 24-hour BP values during monitoring with the SpaceLabs device, whereas sleep BP measurements were similar between devices (**Supplemental Table 1**).

### Participant Experience

Comfort ratings were higher with the ABPM Pro than SpaceLabs device (median 7.0 [IQR, 5.0-8.5] vs 3.5 [2.0-6.5]; P=0.004); 13 participants (65%) reported greater comfort with ABPM Pro, 5 (25%) reported similar comfort, and 2 (10%) reported greater comfort with SpaceLabs. ABPM Pro was also associated with lower pain (1.5 [0-3.5] vs 5.0 [0.5-7.0]; P=0.003) and embarrassment (0.5 [0-3.5] vs 3.0 [0-6.0]; P=0.01). Sleep interference, daytime activity interference, noise, and skin irritation did not differ significantly (**Table 3**).

**Table 3.** Participant Experience Ratings Following Ambulatory Blood Pressure Monitoring.

| **Outcome** | **SpaceLabs** | **ABPM Pro** | **P value** |
| --- | --- | --- | --- |
| Comfort <sup>a</sup> | 3.5 (2.0–6.5) | 7.0 (5.0–8.5) | 0.004 |
| Pain <sup>b</sup> | 5.0 (0.5–7.0) | 1.5 (0–3.5) | 0.003 |
| Embarrassment <sup>c</sup> | 3.0 (0–6.0) | 0.5 (0–3.5) | 0.01 |
| Sleep interference <sup>d</sup> | 4.0 (1.5–6.0) | 2.5 (0–6.0) | 0.08 |
| Daytime activity interference <sup>e</sup> | 4.5 (1.5–7.5) | 3.5 (0–5.0) | 0.05 |
| Noise <sup>f</sup> | 1.5 (0–3.5) | 0 (0–4.0) | 0.39 |
| Skin irritation <sup>g</sup> | 0.5 (0–3.0) | 1.0 (0–4.5) | 0.64 |

Values are presented as median (interquartile range). Abbreviations: IQR, interquartile range.
Note: P values were calculated using Wilcoxon signed-rank tests.
<sup>a</sup> Did you find the monitor comfortable to wear? (0 = not at all comfortable, 10 = extremely comfortable)
<sup>b</sup> Did you experience pain from the monitor? (0 = not at all, 10 = extremely)
<sup>c</sup> Did you feel embarrassed wearing the monitor in public? (0 = not at all, 10 = extremely)
<sup>d</sup> Did the monitor interfere with your normal sleeping pattern? (0 = not at all, 10 = extremely)
<sup>e</sup> Did you find the monitor to interfere with your daytime activity? (0 = not at all, 10 = extremely)
<sup>f</sup> Did you find the noise from the monitor to be disturbing? (0 = not at all, 10 = extremely)
<sup>g</sup> Did you experience skin irritation from wearing the monitor? (0 = not at all, 10 = extremely)

## Discussion

In this randomized crossover study, both ABPM devices generated recordings that met predefined quality criteria. Participants reported significantly greater comfort, less pain and embarrassment while wearing the ABPM Pro device compared with the SpaceLabs device. To our knowledge, this is the first randomized study directly comparing these two ABPM devices.

Our study extends prior work evaluating participant experience with ABPM. Prior data show that ABPM commonly causes discomfort and sleep disturbance.^4-7^ For example, in a randomized study of 365 adults comparing 3 traditional ABPM devices, Nwankwo et al. found that most participants achieved adequate recordings but more than one-half reported sleep disturbance during monitoring.^5^ Although comfort, pain, and embarrassment favored the ABPM Pro, sleep interference did not differ, suggesting that different aspects of participant experience may be influenced by distinct device characteristics.

These findings are relevant to ongoing efforts to improve implementation of ABPM in clinical practice and align with priorities identified by a recent NHLBI workshop report highlighting the importance of evaluating patient acceptability of novel validated BP technologies.^2^ Traditional oscillometric ABPM devices typically require a cuff connected by tubing to an external monitor, whereas the ABPM Pro device uses a compact tubeless cuff-integrated design. Although this study was not designed to identify mechanisms underlying participant preference, the more favorable experience with the ABPM Pro suggests that device design may influence the acceptability of ambulatory BP monitoring.

This study has several strengths, including the randomized crossover design, enabling within-participant comparison of device experience, use of actigraphy-defined sleep periods, and direct comparison of two validated ABPM devices. Several limitations should also be considered. The sample size was small, and the study was conducted at a single center, limiting generalizability. In addition, participants wore the devices on separate days, making it difficult to distinguish whether the device-related differences in awake and 24-hour BP reflect normal day-to-day variability in BP or device-specific measurement characteristics. Accordingly, these BP findings should be considered hypothesis-generating and require confirmation in larger studies specifically designed to assess measurement comparability.

In conclusion, both the SpaceLabs 90227 ABPM and SOMNOmedics ABPM Pro devices produced recordings that met predefined ambulatory BP quality criteria. The ABPM Pro was associated with greater comfort and less pain and embarrassment. These findings support larger comparative studies to determine whether improvements in ABPM device design can enhance patient acceptability and reduce barriers to ABPM implementation in clinical practice.

## Data Availability

All data produced in the present study are available upon reasonable request to the authors

## References

1. Jones DW, Ferdinand KC, Taler SJ, Johnson HM, Shimbo D, Abdalla M, et al. 2025 AHA/ACC/AANP/AAPA/ABC/ACCP/ACPM/AGS/AMA/ASPC/NMA/PCNA/SGIM Guideline for the Prevention, Detection, Evaluation and Management of High Blood Pressure in Adults: A Report of the American College of Cardiology/American Heart Association Joint Committee on Clinical Practice Guidelines. Circulation. 2025;0(0),

2. Abdalla M, Juraschek SP, Biaggioni I, Bowling CB, Balijepalli RC, Brady TM, et al. Blood Pressure Assessment Across the Lifespan: Improving Clinical Research and Clinical Practice: A National Heart, Lung, and Blood Institute Workshop Report. JACC. 2026;87(18):2524–42,

3. Kronish IM, Kent S, Moise N, Shimbo D, Safford MM, Kynerd RE, et al. Barriers to conducting ambulatory and home blood pressure monitoring during hypertension screening in the United States. J Am Soc Hypertens. 2017;11(9):573–80.PMC:5595651

4. Carter EJ, Moise N, Alcantara C, Sullivan AM, Kronish IM. Patient Barriers and Facilitators to Ambulatory and Home Blood Pressure Monitoring: A Qualitative Study. Am J Hypertens. 2018;31(8):919–27.PMC7190918

5. Nwankwo T, Coleman King SM, Ostchega Y, Zhang G, Loustalot F, Gillespie C, et al. Comparison of 3 Devices for 24-Hour Ambulatory Blood Pressure Monitoring in a Nonclinical Environment Through a Randomized Trial. Am J Hypertens. 2020;33(11):1021–9.PMC:7641984

6. van der Steen MS, Lenders JW, Thien T. Side effects of ambulatory blood pressure monitoring. Blood Press Monit. 2005;10(3):151–5,

7. Gaffey AE, Schwartz JE, Harris KM, Hall MH, Burg MM. Effects of ambulatory blood pressure monitoring on sleep in healthy, normotensive men and women. Blood Press Monit. 2021;26(2):93–101.PMC7933045

8. Roth B, Bothe TL, Patzak A, Pilz N. Validation of the ABPMpro ambulatory blood pressure monitor in the general population according to AAMI/ESH/ISO Universal Standard (ISO 81060–2:2018). Blood Press Monit. 2023;28(3):158–62,

9. ABPMPro FDA Clearance Letter 2023 [cited 2026 June 3]. Available from: https://www.accessdata.fda.gov/cdrh_docs/pdf23/K231104.pdf.

10. de Greeff A, Shennan AH. Validation of the Spacelabs 90227 OnTrak device according to the European and British Hypertension Societies as well as the American protocols. Blood Press Monit. 2020;25(2):110–4,

11. Stergiou GS, Palatini P, Parati G, O’Brien E, Januszewicz A, Lurbe E, et al. 2021 European Society of Hypertension practice guidelines for office and out-of-office blood pressure measurement. J Hypertens. 2021;39(7):1293–302,

12. McEvoy JW, McCarthy CP, Bruno RM, Brouwers S, Canavan MD, Ceconi C, et al. 2024 ESC Guidelines for the management of elevated blood pressure and hypertension. Eur Heart J. 2024;45(38):3912–4018,

13. Octavio JA, Contreras J, Amair P, Octavio B, Fabiano D, Moleiro F, et al. Time-weighted vs. conventional quantification of 24-h average systolic and diastolic ambulatory blood pressures. J Hypertens. 2010;28(3):459–64,

14. Abdalla M, Sakhuja S, Akinyelure OP, Thomas SJ, Schwartz JE, Lewis CE, et al. The association of actigraphy-assessed sleep duration with sleep blood pressure, nocturnal hypertension, and nondipping blood pressure: the coronary artery risk development in young adults (CARDIA) study. J Hypertens. 2021;39(12):2478–87.PMC:8571489

