## Supplemental Materials for "Participant Experience with the SpaceLabs 90227 ABPM and SOMNOmedics ABPM Pro Devices"

### Table of Contents

| A. **Supplemental Methods.** | Pages 2-3 |
| --- | --- |
| B. **Supplemental Results.** | Page 3 |
| C. **Supplemental References**. | Page 4 |
| D. **Supplemental Table 1.** Ambulatory Blood Pressure Measurements by Device. | Page 5 |
| E. **Supplemental Figure 1.** Timeline of Study Visits. | Page 6 |

**Supplemental Methods**

Participants were recruited from the community surrounding Columbia University Irving Medical Center and underwent screening to determine eligibility. Eligible participants were English-speaking adults aged 18 years or older. Exclusion criteria included end-stage renal disease requiring dialysis, left ventricular continuous-flow assist devices, pregnancy or plans to become pregnant during the study period, arm circumference >46 cm, lymphedema or inability to wear an ambulatory blood pressure monitor (ABPM) or wrist actigraphy devices, inability to complete study questionnaires, or lack of access to a telephone with text messaging capabilities.

*Study Design and Procedures*

Participants completed three study visits and two separate 24-hour ambulatory blood pressure (BP) monitoring periods during a 7-day monitoring period (**Supplemental Figure 1**). Participants were randomized in a 1:1 ratio to one of two monitoring sequences: (1) SpaceLabs followed by ABPM Pro or (2) ABPM Pro followed by SpaceLabs.

At Visit 1, informed consent was obtained and sociodemographic, medical history, sleep, and anthropometric data were collected. Participants completed baseline sleep questionnaires, including the Pittsburgh Sleep Quality Index (PSQI)^1^ and Insomnia Severity Index (ISI).^2^ Following a 5-minute seated rest period, three office BP measurements were obtained using a validated automated oscillometric device (Omron HEM-907XL) with at least 1 minute between measurements. The average of the three measurements was used for analysis. Office hypertension was defined as mean systolic BP ≥130 mmHg or mean diastolic BP ≥80 mmHg. Participants were then fitted with a wrist actigraphy device (Actiwatch 2 Philips Respironics, Andover, MA) and the first randomly assigned ABPM. Participants were instructed to continue their usual daily activities and maintain their normal sleep schedules while wearing the devices. Participants also completed sleep diaries documenting sleep and wake times, including daytime naps. Participants wore the actigraphy device for the 7-day monitoring period. Sleep and wake periods were determined using actigraphy supplemented by participant sleep diaries.

Following completion of the first 24-hour monitoring period, participants returned to the study center for Visit 2. Office BP measurements were repeated, ABPM data were downloaded, and participants completed a study-specific questionnaire assessing their experience wearing the device. Participants were then fitted with the alternate ABPM according to the randomized crossover sequence. After completing the second 24-hour monitoring period, participants completed a second device experience questionnaire and data from the actigraphy device were downloaded. All participants completed both ABPM monitoring periods and actigraphy and were included in the analysis.

**Supplemental Results**

*Office BP Readings*

There were no significant differences in office BP between visits. Mean SBP ± standard deviation (SD) was 117.0 mmHg ± 14.2 mmHg at visit 1 versus 115.5 ± 13.2 mmHg at visit 2 (p = 0.25). Mean DBP was 76.7 ± 13.4 mmHg at visit 1 versus 75.8 ± 12.8 mmHg at visit 2 (p = 0.17).

**Supplemental References**

1.	Buysse DJ, Reynolds CF, 3rd, Monk TH, Hoch CC, Yeager AL, Kupfer DJ. Quantification of subjective sleep quality in healthy elderly men and women using the Pittsburgh Sleep Quality Index (PSQI). Sleep. 1991;14(4):331–8,

2.	Bastien CH, Vallieres A, Morin CM. Validation of the Insomnia Severity Index as an outcome measure for insomnia research. Sleep Med. 2001;2(4):297–307,

**Supplemental Table 1**. Ambulatory Blood Pressure Measurements by Device.

| **Outcome** | **SpaceLabs Mean ± SD (mmHg)** | **ABPM Pro Mean ± SD (mmHg)** | **P value** |
| --- | --- | --- | --- |
| Awake systolic BP (mmHg) | 124.1 ± 16.2 | 118.3 ± 14.8 | 0.007 |
| Awake diastolic BP (mmHg) | 78.0 ± 12.7 | 74.2 ± 12.3 | 0.008 |
| Sleep systolic BP (mmHg) | 107.3 ± 11.7 | 109.5 ± 14.7 | 0.41 |
| Sleep diastolic BP (mmHg) | 63.3 ± 9.6 | 63.3 ± 12.4 | 0.97 |
| 24-hour systolic BP (mmHg) | 119.2 ± 14.4 | 115.7 ± 14.3 | 0.036 |
| 24-hour diastolic BP (mmHg) | 73.8 ± 11.7 | 71.0 ± 11.9 | 0.005 |

Abbreviations: BP, blood pressure; SD, standard deviation.

Note: P values were calculated using paired t-tests.

**Supplemental Figure 1**. Timeline of Study Visits.

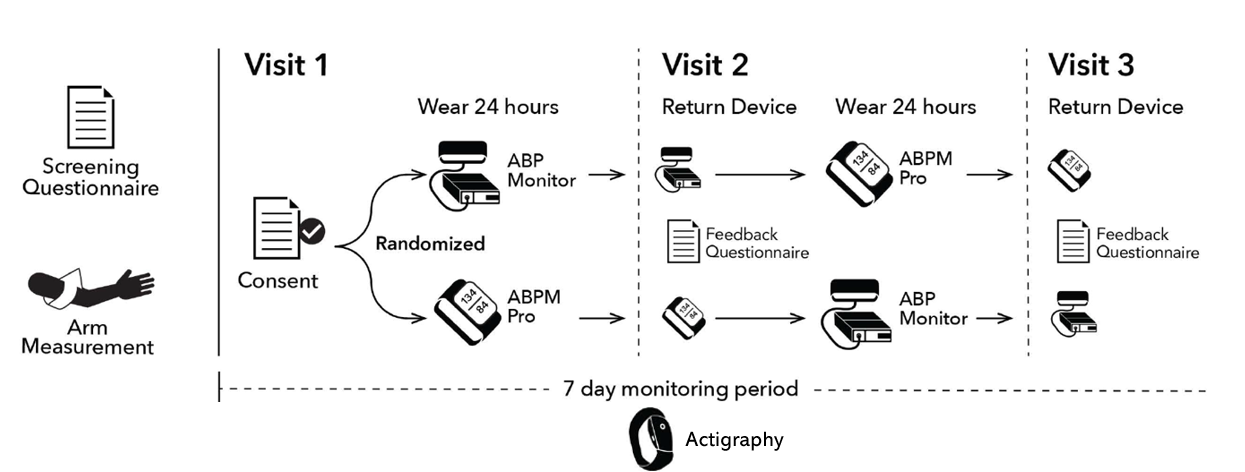
